# A Differential DNA Methylome Signature of Pulmonary Immune Cells from Individuals Converting to Latent Tuberculosis Infection

**DOI:** 10.1101/2021.03.16.21253729

**Authors:** Lovisa Karlsson, Jyotirmoy Das, Moa Nilsson, Amanda Tyrén, Isabelle Pehrson, Nina Idh, Jakob Paues, Cesar Ugarte-Gil, Melissa Méndez-Aranda, Maria Lerm

## Abstract

Tuberculosis (TB), caused by *Mycobacterium tuberculosis*, spreads via aerosols and the first encounter with the immune system is with the pulmonary resident immune cells. The role of epigenetic regulations through DNA methylation in the immune cells is emerging. We have previously shown that capacity to kill *M. tuberculosis* is reflected in the DNA methylome. The aim of this study was to investigate epigenetic modifications in the pulmonary immune cells in a cohort of medical students with a previously documented increased risk of TB exposure, longitudinally. Sputum samples containing alveolar macrophages (AMs) and T cells were collected before and after study subjects worked in hospital departments with a high-risk of TB exposure. DNA methylome analysis revealed that a unique DNA methylation profile was present already at inclusion in subjects who developed latent TB during the study. The profile was both reflected in different overall DNA methylation distribution as well as more profound alterations in the methylation status of a unique set of CpG-sites. Over-representation analysis of the DMGs showed enrichment in pathways related to metabolic reprograming of macrophages and T cell migration and IFN-γ production. In conclusion, we identified a unique DNA methylation signature in individuals, while still IGRA-negative and who later developed latent TB. Epigenetic regulation was found in pathways that have previously been reported to be important in TB. Together the study suggests that DNA methylation status of pulmonary immune cells can predict IGRA conversion.

## Introduction

Tuberculosis (TB) is a major global health concern, ranked as one of the top 10 causes of death worldwide and estimated to be responsible for 1.2 million deaths per year ^1^. TB is caused by the facultative intracellular bacteria *Mycobacterium tuberculosis* and one-fourth of the world’s population is estimated to be infected making *M. tuberculosis* is one of the most successful pathogens known. *M. tuberculosis* is air-born and enters the lung where the bacteria are internalized by alveolar macrophages (AMs) through phagocytosis. The “checkpoint model” can be used to describe the following immunological events post infection ^2,3^. At the first checkpoint, to establish an infection, *M. tuberculosis* needs to evade elimination by AMs. *M. tuberculosis* has developed several strategies to manipulate the host immune response to extend its survival in the phagocytes. The pathogen can arrest maturation of the phagolysosome and direct phagocytes to necrosis, which is prerequisite for the bacterium to spread. In some individuals, referred to as *early clearers*, the pathogen is successfully cleared by innate immune mechanisms at this initial stage of infection ^4–6^. If the pathogen is replicating and cannot be cleared by the innate immune system, the second checkpoint is reached; activation of the adaptive immune system. When the infection is controlled by the adaptive immune system, asymptomatic latent TB infection (LTBI) has developed. From this point, there is a 5-10% lifetime risk of progression to active TB (ATB), as a result of inadequate immune control, which is the third checkpoint for *M. tuberculosis* ^7–10^. The Interferon-Gamma Release Assay (IGRA) is an immunological test used to confirm whether a person has been exposed to *M. tuberculosis* based on peripheral T cell release of interferon-γ (IFN-γ) in response to *M. tuberculosis* antigens ^11^. Another method to confirm exposure is the purified protein derivative (PPD) or Tuberculin skin test, where PPD-antigen is injected intradermally to the forearm and the skin reaction, based on T cell-mediated delayed hypersensitivity response against the antigen, is measured ^12^.

The role of epigenetics in TB immune responses is emerging. Several studies have described the concept of developing trained immunity through epigenetic reprogramming, leading to proper orchestration of gene expression upon re-exposure to a pathogen or pathogen-derived products ^3,13,14^. DNA methylation, histone modifications, and regulation of small RNAs are considered as the important players in the regulation of epigenetic modifications ^15^. DNA methylations in CpG-rich promotor regions are generally associated with gene silencing ^16^. We and others have described the reprogramming of DNA methylation patterns in peripheral blood mononuclear cells (PBMCs) after exposure to live attenuated *Mycobacterium bovis* through the *Bacillus Calmette Guérin* (BCG) vaccination ^17,18^. Further, we have demonstrated that differences in these DNA methylation patterns affect the efficacy of the macrophages to kill *M. tuberculosis in vitro* ^19^. A wealth of literature focus on adaptive immune responses in peripheral blood, but since *M. tuberculosis* primarily infects the lung, it would be more relevant to address pulmonary immunity, including alveolar T cells and AMs ^20,21^.

Here, we have increased our focus on the pulmonary-resident immune cells and further investigated DNA methylomes of AMs and alveolar T cells in a cohort of medical students with a previously reported increased risk of *M. tuberculosis* exposure ^22^. The aim of the study was to investigate epigenetic modifications in the pulmonary immune pre- and post-TB exposure in a natural setting. We hypothesized that this recent exposure to *M. tuberculosis* in the lung compartment would induce epigenetic alterations in AMs and alveolar T cells. By sputum induction AMs and alveolar T cells were isolated ^23^. Using reduced representation of bisulfite sequencing (RRBS) the DNA methylome was investigated and we identified a different global DNA methylation profile in pulmonary immune cells of the study subjects that developed latent TB infection during the study. Notably, this global DNA methylation profile was identified in the immune cells already before a latent TB infection could be detected with the IGRA test.

## Results

### Study design and cohort

Medical students were invited to participate in the study and donated sputum samples before (referred here as 0 months) and after (referred here as 6 months) clinical rotations in departments with a high-risk of *M. tuberculosis* exposure. A schematic overview of the study design is represented in Figure 1. Demographic information of the study subjects and IGRA results are shown in Table 1. One study subject was borderline IGRA-positive ^24^ (IGRA_pos_) already at inclusion (0 months) (Table S1). Two study subjects developed latent TB infection during the study as demonstrated with a positive IGRA test at follow-up (6 months, referred to as IGRA converters in the following text) (Table S1).

**Table 1.** Demographics of study subjects. A total of 15 participants were included in the study. IGRA test confirmed IGRA conversion in two study subjects. ^⍰^ shows the range of the data.

| Characteristics | Participants (N=15) |
| --- | --- |
| Sex (male and female) | 8 / 7 |
| Age (years) | 22.7 (21-29) <sup>a</sup> |
| Weight (kg) | 70.5 (48.5-101) <sup>a</sup> |
| Height (cm) | 168.8 (159-182) <sup>a</sup> |
| BMI (kg/m<sup>2</sup>) | 24.6 (17.2-34.5) <sup>a</sup> |
| IGRA result 0 months (negative and positive) | 14 / 1 |
| IGRA result 6 months (negative and positive) | 12 / 3 |

**Figure 1.**
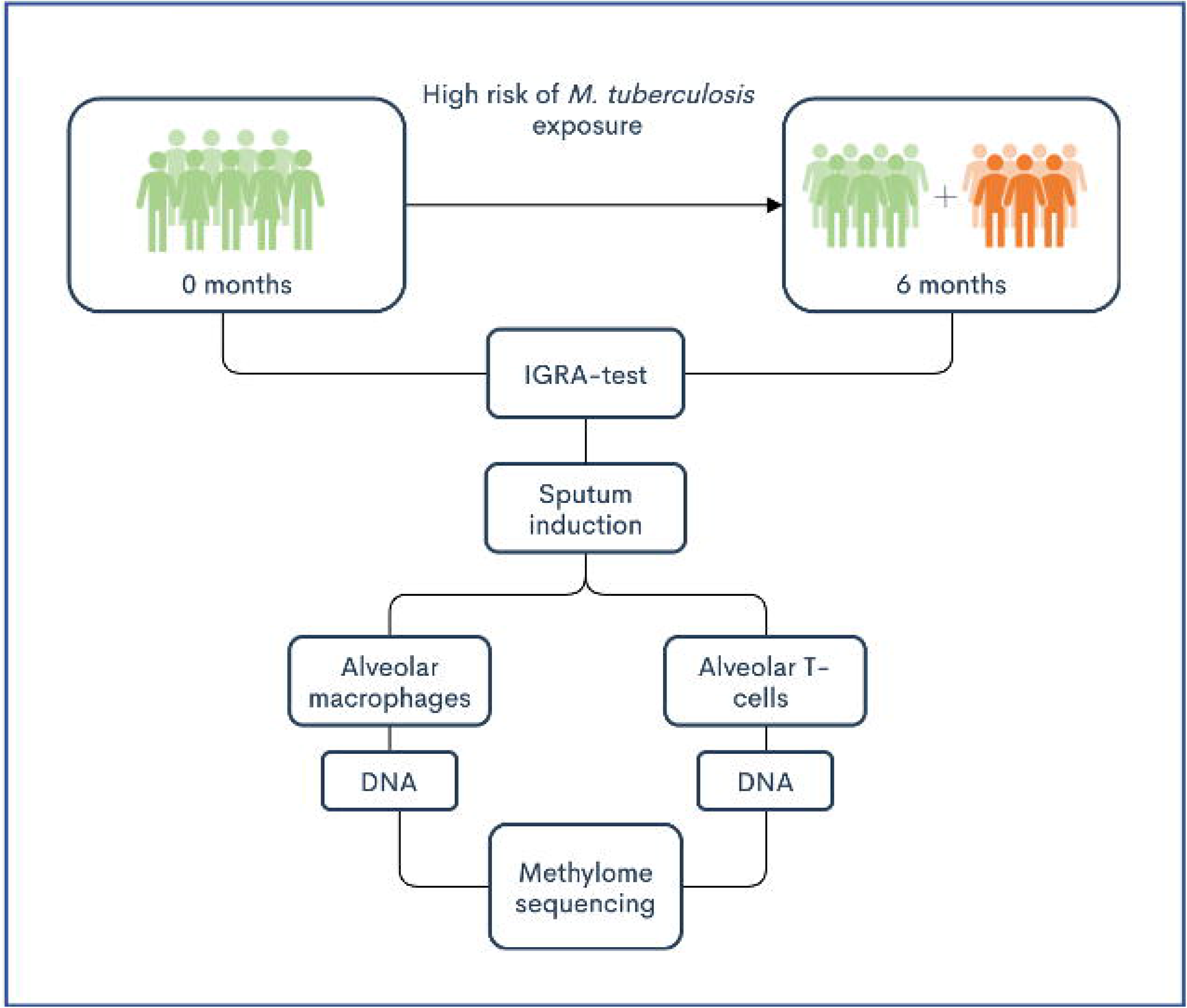
Flow chart of the study design. Study subjects donated sputum and blood samples at 0 and 6 months, which corresponded to before and after clinical rotations at departments with high-risk of *M. tuberculosis* exposure at hospitals in Lima, Peru. IGRA tests were taken to confirm TB-infection. AMs and alveolar T cells were isolated from sputum samples. DNA was extracted and the samples were sequenced with reduced representation bisulfite sequencing (RRBS) for methylation analysis.

### DNA methylation patterns in alveolar macrophages and T cells before and after TB exposure distinguish IGRA converters

To investigate the epigenetic changes in the pulmonary immune cells over time, we isolated DNA from the AMs and alveolar T cells collected at 0 and 6 months and performed Genome-wide Reduced Representation Bisulfite sequencing (RRBS). After filtering the data as described in the method section, we identified a total of 1186 CpG-sites with ≥5 reads in the 19 samples from the AMs and 404 CpG-sites in the 19 samples from the alveolar T cells. To get an overview of the global DNA methylation distribution in the pulmonary immune cells, we made density plots of the M-values (the log_2_ ratio of the intensities of methylated versus unmethylated CpG-sites) obtained from CpG-sites in the AMs (1186 CpG-sites) and the alveolar T cells (404 CpG-sites), shown in Figure 2A,B. The density plot showed a homogenous distribution of DNA methylation in all samples from the IGRA_neg_ study subjects collected at both at 0 and 6 months. The sample from one study subject, that was IGRA_pos_ at inclusion, followed the same global DNA methylation distribution. Whereas in the IGRA-converting study subjects, we identified a different global DNA methylation profile with more hyper- and hypomethylated CpG-sites as well as different peak densities. Notably, the sample collected at 6 months (after IGRA conversion) displayed a similar DNA methylation profile as the sample collected at inclusion.

**Figure 2.**
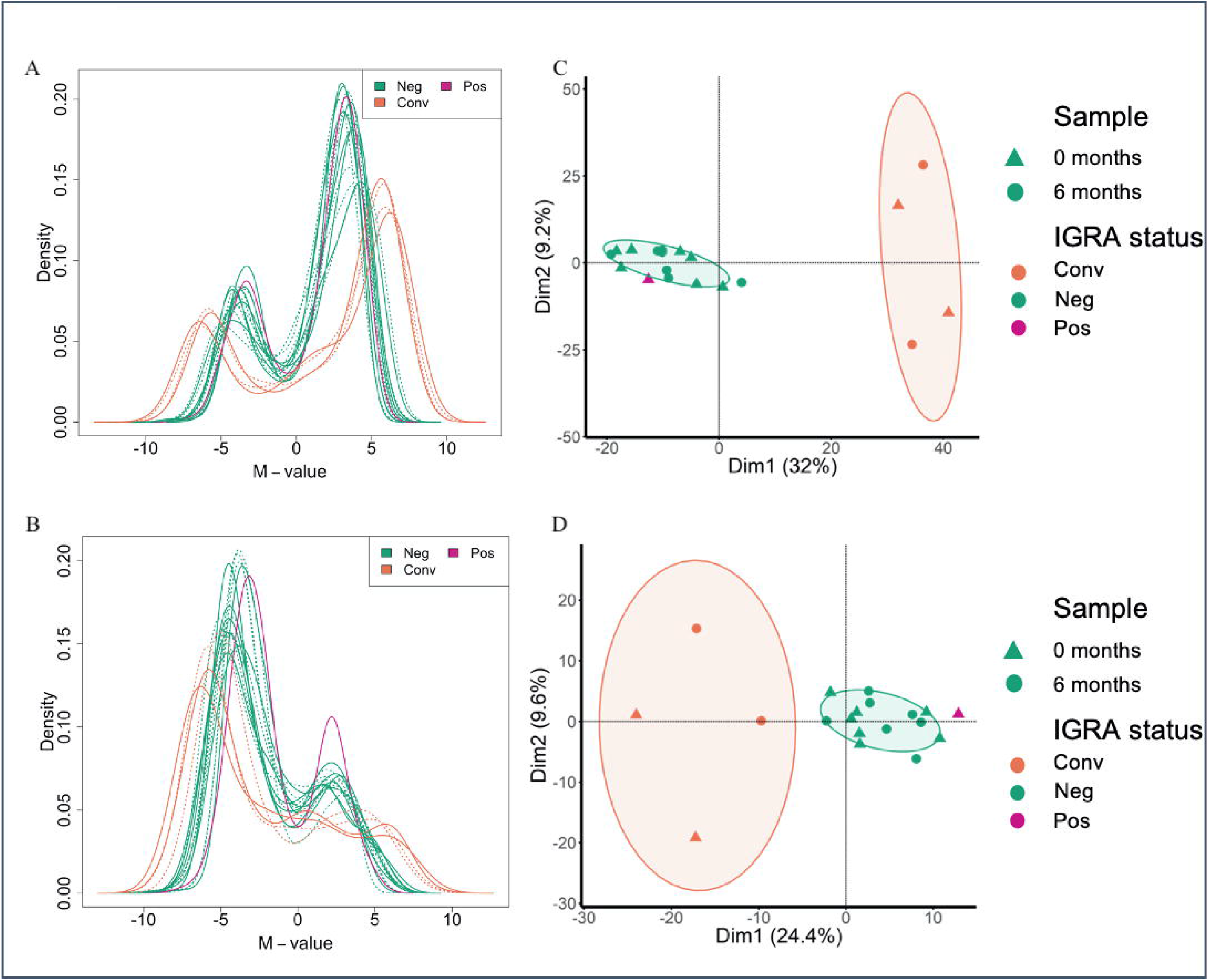
Unique distribution of genomic DNA methylation discriminates IGRA-converting individuals. Density plots of the distribution of M-values from CpG-sites identified in **A**. AMs and **B**. alveolar T cells. Full line represents samples collected at 0 months and dotted lines samples collected at 6 months. The Principal Component Analysis (PCA) of the methylation data from **C**. AMs and **D**. alveolar T cells. The ellipses represent the 70% confidence interval (C.I.) in the dataset.

The results coincided in both the AMs and the alveolar T cells. To further explore the data, we performed a principal component analysis (PCA) which revealed a distinct group formation from the IGRA converters (based on the PC1 (Dim1) with 70% C.I.), including both the samples collected before and after the IGRA conversion (Figure 2C,D). The IGRA_pos_ individual on the other hand clustered with the data obtained from IGRA_neg_ study subjects. We proceeded with a hierarchical clustering analysis by applying the Euclidean similarity/dissimilarity matrix calculation using the M-value (Figure 3A,B). In line with the PCA results, the data from the IGRA converters formed a separate cluster, including both the sample collected before and after IGRA conversion. The same cluster separation was found in both the samples from the AMs and from the alveolar T cells. To visualize the methylation status in the 1186 and 404 CpG-sites from the AMs and T cells respectively, we created heatmaps (Figure 4A,B), demonstrating a clear difference in the overall methylation between IGRA converters and IGRA_neg_ study subjects.

**Figure 3.**
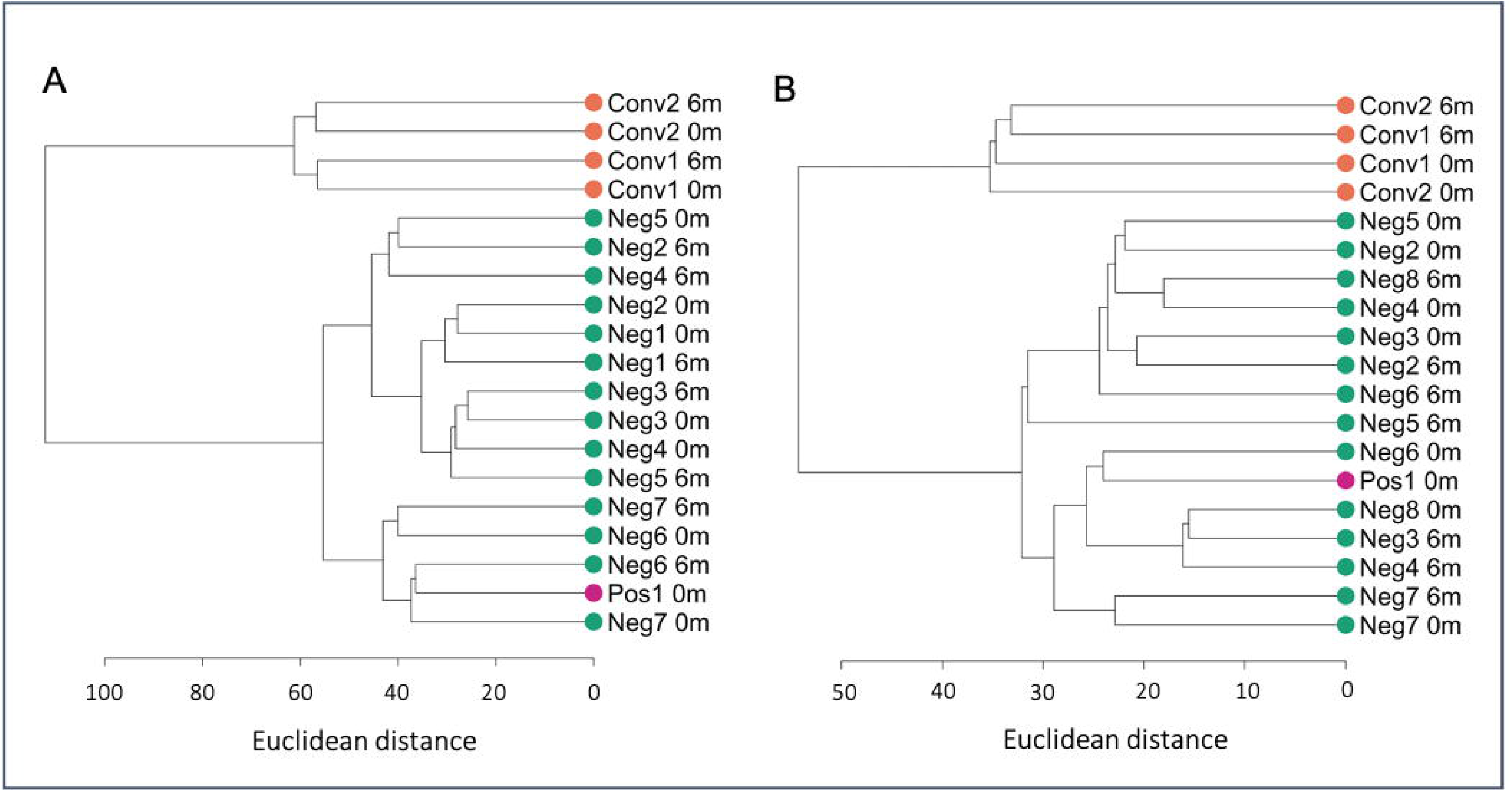
Hierarchical clustering analysis separating IGRA-converting individuals. **A,B**. Unsupervised hierarchical clustering dendrogram applying the Euclidian distance matrix calculation and Ward D2 method. The dendrograms show the clustering of methylome data from **A**. AMs. and **B**. alveolar T cells. The scale represents the Euclidean distance.

**Figure 4.**
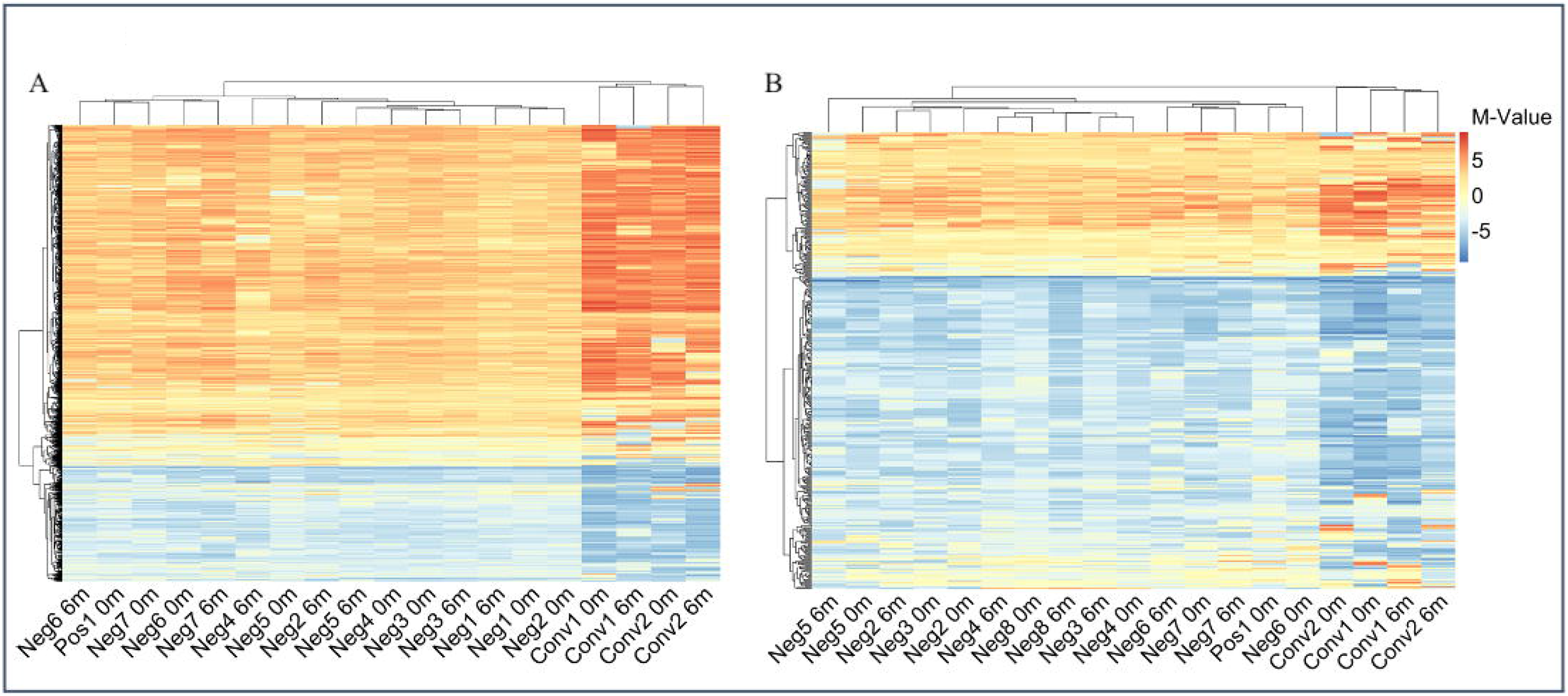
Heatmaps reveal a different overall DNA methylation pattern in IGRA-converting study subjects. **A**. Heatmap of all CpG-sites (1186) identified in the AMs of each study subject. **B**. Heatmap of all CpG-sites (404) identified from the alveolar T cells. The color bar represents the M-value scale ranging from −10 (blue) to 10 (red).

## High number of strong DMCs identified in IGRA-converting individuals

To further explore the epigenetic reprogramming that occurred between the two time points, 0 and 6 months, we identified the differentially methylated CpG-sites (DMCs) with the strict filtering criteria (|log_2_ Fold Change (log_2_FC)| > 5 and Benjamini-Hochberg (BH) corrected *p*-value < 0.01) in each study subject. The DMCs were divided into three different cutoff levels: |log_2_FC| 10, 13 and 15 hyper- or hypomethylation in order to understand the distribution of DMCs based on the level of change (Figure 5A,B). A high number of DMCs with a |log_2_FC| >15 was identified in the IGRA converters, in both cell types. To normalize the results to account for the different number of total DMCs (|log_2_FC| > 5, BH adj. *p-*value < 0.01) identified we looked at the percentage of DMCs with a |log_2_FC| > 15 (Figure 5C).

**Figure 5.**
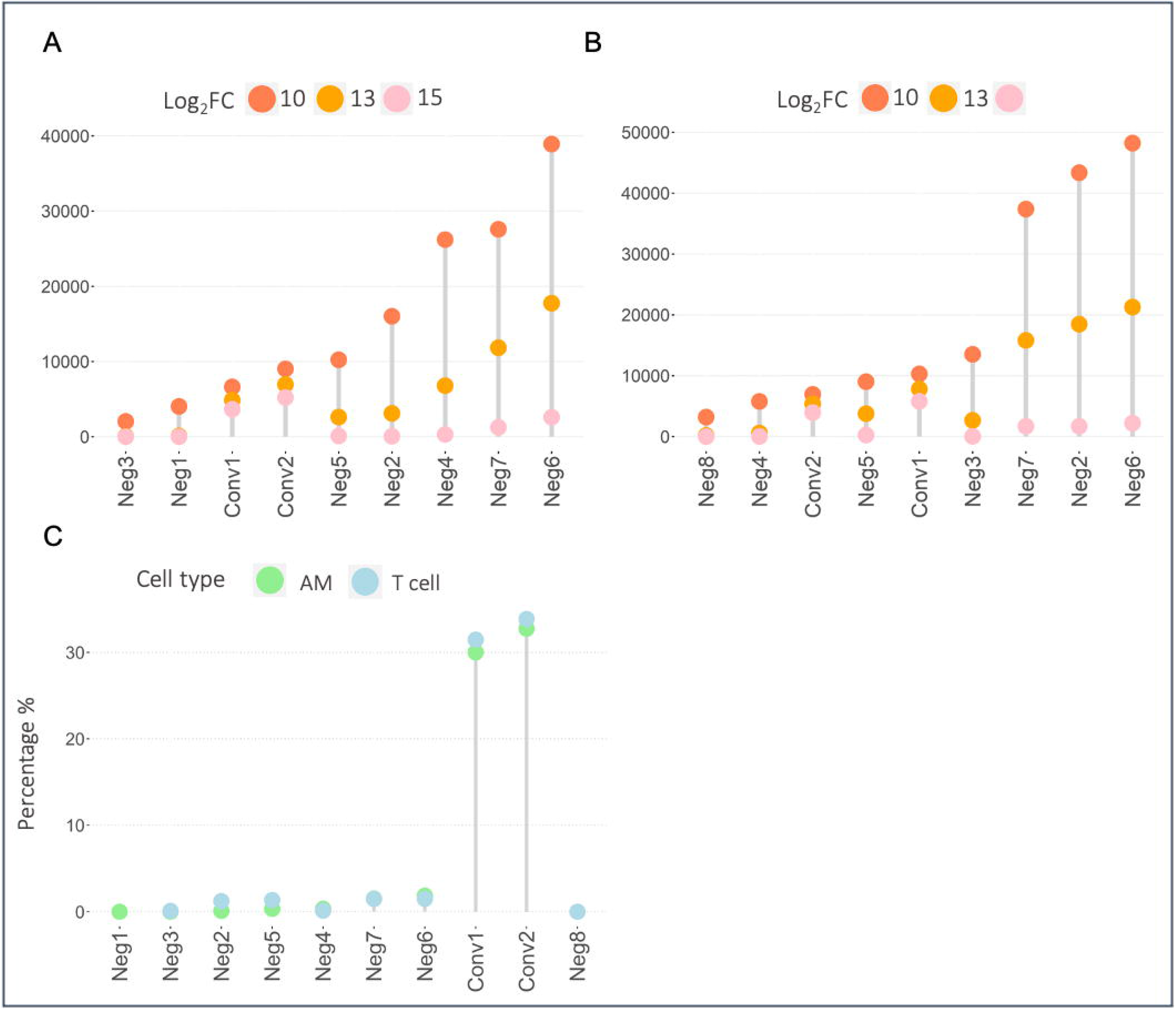
Profound DNA methylome alterations identified in IGRA-converting study subjects. Lollipop plots showing the number of differentially methylated CpG-sites (DMCs) identified in the **A**. AMs and **B**. alveolar T cells of each study subject. C. The percentage of DMCs with a |log_2_FC| >15 compared to the total number of DMCs (|log_2_FC| > 5, adj *p*-value > 0.01) identified in each study subject AMs and T cells.

### IGRA-converting individuals undergo similar epigenetic changes in DNA methylation

The DMCs were annotated to the official Gene Symbols further analysis and is referred to as DMGs in the following text. To identify the overlap in the DMGs with a |log_2_FC| >15 between the study subjects, a Venn analysis was performed and presented in an UpSet plot. The IGRA converters shared the largest intersection in both cell types, 452 DMGs in the AMs (Figure 6A) and 471 DMGs in the T cells (Figure S1). 32 DMGs overlapped between the AMs and T cells. To filter out unspecific changes we selected the DMGs that changed in the same direction in both IGRA converters. We identified 280 (128 DMGs were hypermethylated and 152 hypomethylated) and 281 (159 DMGs were hypermethylated and 122 hypomethylated) DMGs that became hypo- or hypermethylated, in both IGRA converters in the AMs and in the T cells respectively.

**Figure 6.**
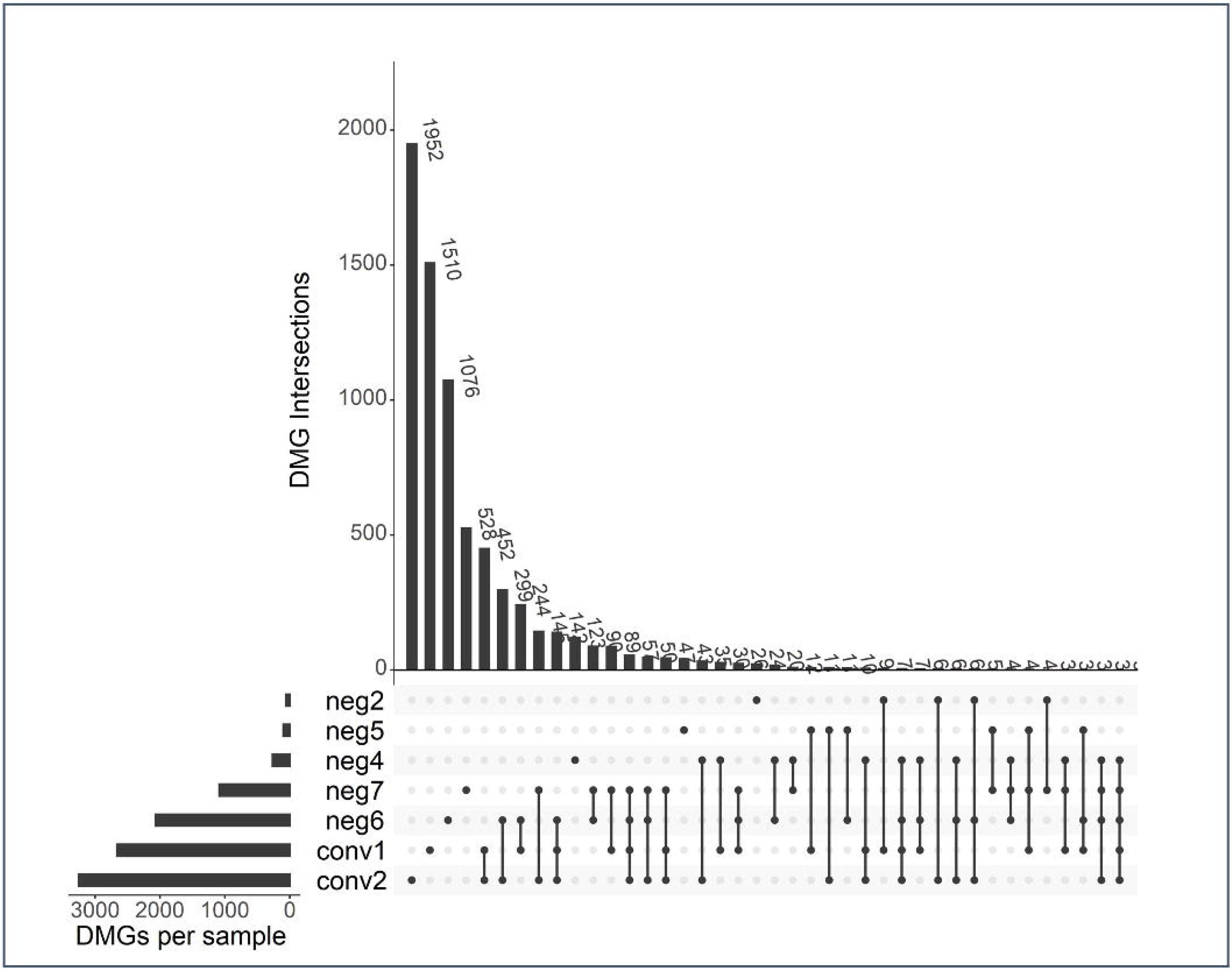
IGRA converters undergo unique DNA methylation changes during IGRA conversion. UpSet plot showing the intersects of the DMGs identified in each study subjects AMs.

### IGRA converters’ DMGs are over-represented in pathways related to metabolic reprograming, T cell migration and IF N-γ production

In an over-representation analysis (ORA) ^25^ using the PANTHER database ^26^ the identified DMGs from the AMs unique to the IGRA converters were shown to be overrepresented in the Pentose phosphate pathway and the Ras pathway (Table 2). In the ORA of the DMGs from the T cells unique to the IGRA converters we identified muscarinic acetylcholine receptors 1-4 pathway and β1- and β2 adrenergic receptor signaling pathway (Table 3).

**Table 2.** ORA of DMG intersect from IGRA converter’s AMs. ORA using Panther pathways of the DMG intersect identified in the AMs of the IGRA converters.

| Gene Set | Description | Ratio | p-value | FDR |
| --- | --- | --- | --- | --- |
| P02762 | Pentose Phosphate Pathway | 20.981 | 0.004 | 0.41 |
| P04393 | Ras Pathway | 3.596 | 0.049 | 1 |

**Table 3.**
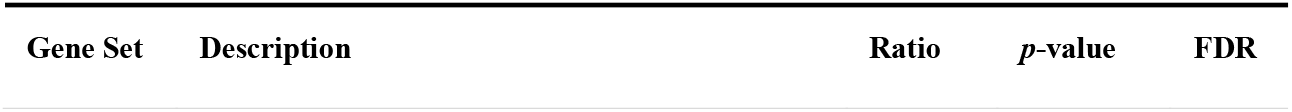
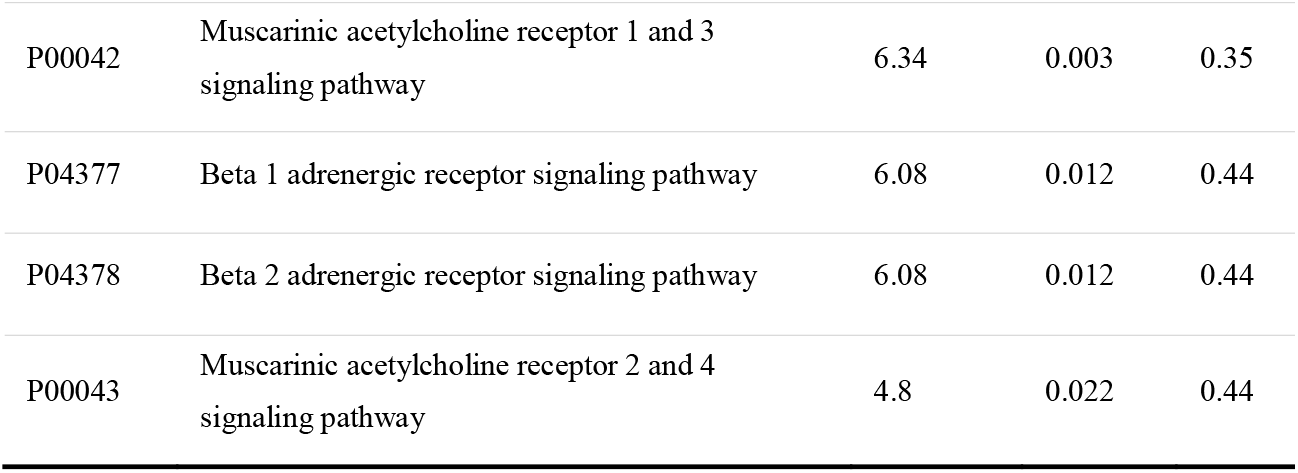
ORA of DMG intersect from IGRA converter’s alveolar T cells. ORA of the DMG intersect identified in the T cells of the IGRA converters.

## Discussion

In this study we investigated the genome-wide DNA methylation in the pulmonary immune cells in individuals at high risk of exposure to *M. tuberculosis*. DNA methylation has been shown to play an important role in the innate immune systems response to mycobacteria by us and others ^17,27–30^, however, it has not yet been longitudinally studied in healthy individuals at risk of TB exposure. Two of the study subjects in this study developed latent TB in the course of sample collection as demonstrated by IGRA conversion and our analyses identified a distinct genome-wide DNA methylation profile separating these two individuals from those who remained IGRA_neg_ throughout the study. This DNA methylation profile, which, contrary to our expectation, was present already at inclusion, was characterized by more hypo- and hypermethylated CpG-sites. Intriguingly, the subject diagnosed with latent TB at inclusion, did not cluster with the IGRA converters. From these observations, we propose the following hypotheses: i) the IGRA converters were exposed to TB via the airways already before inclusion and the exposure had not yet developed into circulating T cell memory (resulting in a negative IGRA test) but had caused reprogramming of the DNA methylome of AMs; ii) a different, *inherent* genomic DNA methylation profile as observed in the IGRA converters predispose for IGRA conversion in a high-endemic setting.

According the first hypothesis, the observed DNA methylation pattern represents a normal, protective response to TB exposure (as part of the first ‘checkpoint’), which however was breached by the infection and therefore urged the host to induce an adaptive immune response (the second ‘checkpoint’) ^2,3^. If the second hypothesis applies, a limited capacity for inducing a host-protective epigenetic reprogramming could cause susceptibility and predisposition for latent TB infections. There is a large heterogeneity in the human susceptibility to develop latent or active TB upon exposure and there is limited knowledge in the determining factors ^31^, but larger studies could investigate the correlation between DNA methylation patterns and TB susceptibility. Alternatively, the distinct DNA methylation pattern observed could also be affected by *M. tuberculosis* manipulating the host cells epigenome ^28,32,33^.

The two IGRA converters underwent similar epigenetic changes between the timepoints, during conversion to IGRA positivity. There was a large overlap in DMGs in the IGRA converters in both cell types. The DMG intersect unique to the IGRA converters AMs showed over-representation in the pentose phosphate pathway which has previously been described to be critical in *M. tuberculosis* infected macrophages ^34,35^. Infected macrophages become dependent on both glycolysis and the pentose phosphate pathway to fulfill the metabolic and bioenergetic requirements for production of cytokines, chemokines and reactive oxygen species (ROS) ^34–36^. The DMGs unique to the IGRA converters’ T cells showed over-representation in the pathway for β-adrenergic and muscarinic acetylcholine receptor signaling. A functional role between β2 adrenergic receptor (β2AR) activation and IFN-γ and Tumor necrosis factor (TNF) production in T helper 1 (Th1) cells have been established in several studies, activation of β2AR by the neurotransmitter norepinephrine (NE) decreases the Th1 cells IFN-γ expression ^37–40^. Polymorphisms in the β2AR have also been reported to be associated with TB ^41^. The cholinergic system also has a functional role in the T cells. T cells express both muscarinic and nicotinic acetylcholine (ACh) receptors and expression of the enzyme choline acetyltransferase (ChAT) is induced in T cells during infection, T cell derived ACh is involved in T cells migration to tissues ^42–44^.

One study subject was borderline IGRA positive at inclusion and this subject followed the same DNA methylation distribution and was clustering with the IGRA_neg_ study subjects. The kinetics of the IFN-γ-reaction in relation to point of infection is not elucidated and after TB treatment the peripheral T cell memory will persist for up to 15 months ^45,46^. According to our analysis, the epigenome of the pulmonary immune cells in this individual was more similar to that of healthy individuals and this suggests the possibility that this individual became IGRA positive a long time ago and that the epigenome profile in the pulmonary immune cells has returned to baseline.

If the DNA methylation profile observed in the IGRA converters is an early result of *M. tuberculosis* infection, the finding could have implications as a diagnostic tool for identification of individuals that are developing latent TB infection before any of the currently available diagnostic methods. However, if the patterns we identified is not a result of bacterial infection but rather an epigenomic dysregulation predisposing individuals to convert upon exposure, this global DNA methylation pattern could have implications in identifying risk-groups who are more prone to convert upon exposure.

We recognize that this study was performed on a small number of study subjects and further investigation in larger cohorts is needed to elucidate the results presented here. We also acknowledge the limitations of the study design, we included a cohort of study subjects with risk of *M. tuberculosis* exposure, but no definite measurement of exposure. The observations made with IGRA converters were limited to two study subjects. However, we highlight the significance of investigating pulmonary immune cells and the value of investigating DNA methylomes in the same study subjects, longitudinally, before and after IGRA conversion.

## Methods

### Ethical Statement

Ethical approval was obtained from Universidad Peruana Cayetano Heredia (UPCH) Institutional Review Board N° 103793. All participants signed an informed consent.

### Study cohort and design

This was a prospective study aimed to investigate epigenetic DNA methylome alterations in pulmonary immune cells pre- and post-*M. tuberculosis* exposure in a natural setting. We enrolled medical students *(n= 15)* in the fifth or sixth year of medical school at UPCH. The students donated sputum and blood samples before (0 months) and after (6 months) they had clinical rotations in high-risk departments of *M. tuberculosis* exposure. The department of infectious disease and internal medicine including the emergency department were defined as departments with high-risk of *M. tuberculosis* exposure. Sputum was used to isolate immune cells from the lung and the blood was used for IGRA. At inclusion, the participants filled in an individual case report form with demographic information. Participants also filled in an online questionnaire to collect background information before each session at 0 and 6 moths. The questionnaire was created in the tool Survey & Report, provided by Linköping University. Samples from 10 individuals (collected at 0 and 6 months for IGRA_neg_ (*n= 7*) and IGRA converters (*n= 2*) and at 0 months for IGRA_pos_ (*n= 1*)) resulted in 19 samples that was selected for each cell type (AMs and alveolar T cells), for the sequencing analysis. The sample selection was based on results in the online questionnaire and on sample quality with regards to DNA concentration in the obtained in both AMs and T cells at the two sample collections at 0 and 6 months.

### Interferon gamma releasing assay with QuantiFERON® TB-Gold Plus

At inclusion and follow-up, the participants donated blood samples that were used for IGRA with the QuantiFERON® TB-Gold Plus test (SSI Diagnostica, Hillerød, Denmark) according to the manufacturer’s instructions. Tubes were filled and incubated at 37°C for maximum 24 hours and then analyzed with ELISA.

### Sputum Induction

The sputum induction ^23^ was performed with an eFlow rapid nebulizer (PARI, Hamburg, Germany) filled with a hypertonic saline solution. The solution was prepared by mixing sterile water (Fresenius Kabi, Stockholm, Sweden) with 4% sodium chloride (B. Braun Medical AB, Stockholm, Sweden). The participants inhaled the solution for 9 minutes while simultaneously preforming breathing exercises in accordance with instructions from the lung clinic at Linköping University Hospital. The participants were asked to cough deeply to expectorate sputum from the lungs. The expectorates were collected into sterile 50 ml Falcon tubes (Thermo Fisher Scientific, Waltham, US) after each session. The Falcon tubes were kept on ice. The sputum inductions were performed in three replicates.

### Sputum Processing and CD3 and HLA-DR Positive Cell Isolation

From the sputum samples, within two hours, plugs containing pulmonary immune cells ^23^ were picked and pooled. The plugs were dissolved by adding 0.1% dithiothreitol (DTT) (Thermo Fisher Scientific) mixed in phosphate-buffered saline (PBS) (Gibco, Cambridge, UK). This was added in a volume approximately 4 times the collected sputum volume and then vortexed and placed on a tilter with ice for 20 min. The dissolved sputum sample was then filtered through 50 μm cell strainers (Sigma-Aldrich, Saint Louise, US) into a new 50 ml Falcon tube and centrifuged for 5 min at 380g in 4°C. The supernatant was discarded, and the pellet was resuspended in 500 µl of an isolation buffer containing 500 mM ethylenediaminetetraacetic acid (EDTA), 0.1% fetal bovine serum (FBS), Ca^2+^ and Mg^2+^ free PBS (pH 7.4). First, CD3 positive cells were isolated from the resuspended pellet. 25 µl of Dynabeads CD3 (Invitrogen Dynabeads®, Life Technologies AS, Oslo, Norway) were washed with 800 µl PBS and placed in a DynaMag-2 (Thermo Fisher Scientific) for 1 min. The supernatant was discarded and 800 µl isolation buffer were added two times for additional washing. The beads were then mixed with 500 µl of the cell sample and incubated for 30 min at 4°C while tilting. After incubation, the tube was placed in the DynaMag-2, supernatant was removed to be used to isolate HLA-DR positive cells next. The CD3 positive cells were resuspended in 200 µl PBS.

Secondly, HLA-DR positive cells were isolated. 25 µl Magnetic Pan Mouse IgG Dynabeads(tm) (cat no: 11041, Thermo Fisher) were washed with 800 µl PBS and placed in a DynaMag-2 (Thermo Fisher Scientific) for 1 min. The supernatant was discarded and 800 µl isolation buffer were added two times for additional washing. 5 µl monoclonal HLA-DR antibodies (cat no: 14-9956-82, Thermo Fisher) were added to conjugate the Dynabeads. Tubes were incubated for 40 min, then placed in the DynaMag-2 for 30 seconds and supernatant removed. 800 µl isolation buffer were added two times for washing. The tube was placed in DynaMag-2 for 30 seconds and supernatant removed. The beads were then mixed with 500 µl of the supernatant from the CD3 isolation and incubated for 30 min at 4°C while tilting. After incubation, the tube was placed in the DynaMag-2, supernatant was discarded, and HLA-DR positive cells were resuspended in 200 µl PBS. CD3 positive cells are referred to as alveolar T cells (T cells) and HLA-DR positive cells are referred to as alveolar macrophages (AMs).

### DNA Extraction and Quantification

DNA was extracted from the AMs and T cells with the AllPrep® DNA/RNA Mini Kit (Qiagen, Hilden, Germany) within 4 hours from cell isolation. Concentration of DNA was quantified with a Qubit® 4.0 Fluorometer (Thermo Fisher Scientific), using dsDNA High Sensitivity (HS) Assay Kit (Thermo Fisher Scientific). The measurement was performed according to the manufacturer’s instructions.

### Reduced Representation Bisulfite Sequencing of DNA from AMs and T cells

DNA samples were sequenced with Reduced Representation Bisulfide Sequencing (RRBS) at the Bioinformatics and Expression Analysis (BEA) core facility at Karolinska Institute (KI) with Diagenode’s RRBS. The DNA was enzymatically digested, bisulfite converted, and PCR amplified before ready for Illumina’s HiSeq 2000. The use of the restriction enzyme MspI, which cleaves CCGG from the 5′end, results in shorter sequences to analyze and is therefore cost-effective.

### Data processing and computational analysis

The RAW files (in fastq format) were generated from the RRBS analysis, were quality checked using the fastQC ^47^ (v0.11.9). The sequences were trimmed to remove artificially filled-in cytosines at the 3′ end using the TrimGalore (v.0.6.5) ^48^ with a phred score cutoff of 20 and quality checked again after trimming. The trimmed sequences were aligned with the human reference genome (hg38.13) using Bowtie2 ^49^ and removed the deduplicates using the Bismark v.0.22.3 ^50^. The methylation extractor from Bismark was used to extract the CpG methylation data from the sequences. The SAMtools (v1.7) ^51^ package was used to sort the bam files on CpG-site chromosomal location and converted to SAM files. The methylated and unmethylated CpG counts were extracted and combined using the DMRfinder (v0.3) ^52^ package in R (v4.0.2) ^53^.

To read the Bismark coverage files, the *edgeR* (v3.32.1) ^54,55^ package was used. The CpG-sites located in the X and Y chromosome as well as CpG-sites from mitochondrial DNA were filtered out. CpG-sites with a read coverage < 5 with both methylated and unmethylated reads were removed from the analysis. The M-values were calculated using the log_2_ ratio of the intensities of methylated verses unmethylated CpG-sites (with addition of +2 to each count to avoid logarithms of zeros) ^56^.

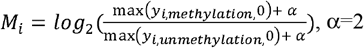

The CpG-sites were annotated using *org*.*Hs*.*eg*.*db* (v3.12) ^57^ and AnnotationDbi (v1.52) ^58^ packages. After filtering and annotating the data, we identified a total of 1186 CpG-sites in the AMs and 404 CpG-sites from T cells that were covered in all samples.

### Statistical analysis

The Principal Component Analysis (PCA) was calculated using the *factoExtra* (v1.0.7) ^59^ and *factoMineR* (v2.4)^60^. The hierarchical clusters was estimated using the *ape* (v5.4-1) ^61^ and *dendextend* (v1.14.0)^62^ by calculating the Euclidian similarity/dissimilarity matrix. For the identification of the individual differentially methylated CpG-sites (DMCs) the EdgeR *lmFit* function ^63^ was used to estimate the counts of the unmethylated (Un) and methylated (Me) reads in the conditions 0 months (A) and 6 months (B) in each individual, calculated as:

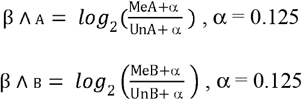

DMCs were defined as CpG-sites with a |log_2_FC| > 5 and significant with the Benjamini-Hochberg (BH) corrected *p*-value < 0.01. The lollipop plots and the upset plots were made using the *ggpubr* (v0.4.0) ^64^ and the *UpSetR* (v1.4.0) ^65,66^ packages. Two IGRA_neg_ study subjects and 1 study subject had no DMCs with a |log_2_FC| > 15 in the AMs and T cells respectively and were therefore excluded from the analysis. The pathway analysis was performed with the DMGs with a |log_2_FC| > 15 and Benjamini-Hochberg (BH) corrected *p*-value < 0.01 unique to the IGRA converters AMs and T cells respectively from the PANTHER database (v16.0) ^26^ using the WEB-based Gene SeT AnaLysis Toolkit (WebGestalt) webserver (v2019) ^25^. The false discovery rate (FDR) in the pathway analysis is BH corrected *p*-values. The significant pathways identified in the AMs and the top 4 most significant pathways identified in the T cells are presented in Table 2.

## Supporting information

Table S1. IGRA test results.

## Data Availability

The datasets will be made available at the final publication.

## Data availability

The datasets will be made available at the final publication.

## Acknowledgements

We thank the Bioinformatics and Expression Analysis core facility (BEA) at Karolinska Institute that preformed the sequencing analysis. We thank the Swedish National Infrastructure for Computing (SNIC) at National Supercomputing Centre (NSC), Linköping University for the computing systems enabling the data handling, partially funded by the Swedish Research Council through grant agreement N° 2018-05973. The work was supported by grants from the Swedish Research Council (Vetenskapsrådet) N° 2018-05973 and N° 2018-04246 and the Consejo Nacional de Ciencia, Tecnología e Innovación Tecnológica CONCYTEC and Cienciactiva N° 106-2018-FONDECYT. J.D is a postdoctoral fellow supported through the Medical Infection and Inflammation Center (MIIC) at Linköping University.

## Author Contributions

N.I, J.P, I.P, C.U-G, M.M-A and M.L wrote the ethical application and conceived the study. I.P, M.N and A.T performed all sample collections. J.D and L.K preformed the bioinformatic analyses. M.L, L.K, J.D and I.P wrote the manuscript. M.M-A and C.U-G supervised the project in Peru.

## Consent for publication

All authors have approved the final version of the manuscript.

## Figure and Table legends

Figure S1. **UpSet plot showing the intersects of the DMGs identified in each study subject’s alveolar T cells**. UpSet plot showing the intersects of the DMGs identified in each study subjects T cells.

