## Supplementary figures and images for "A Differential DNA Methylome Signature of Pulmonary Immune Cells from Individuals Converting to Latent Tuberculosis Infection"

### Supplemental Data 1

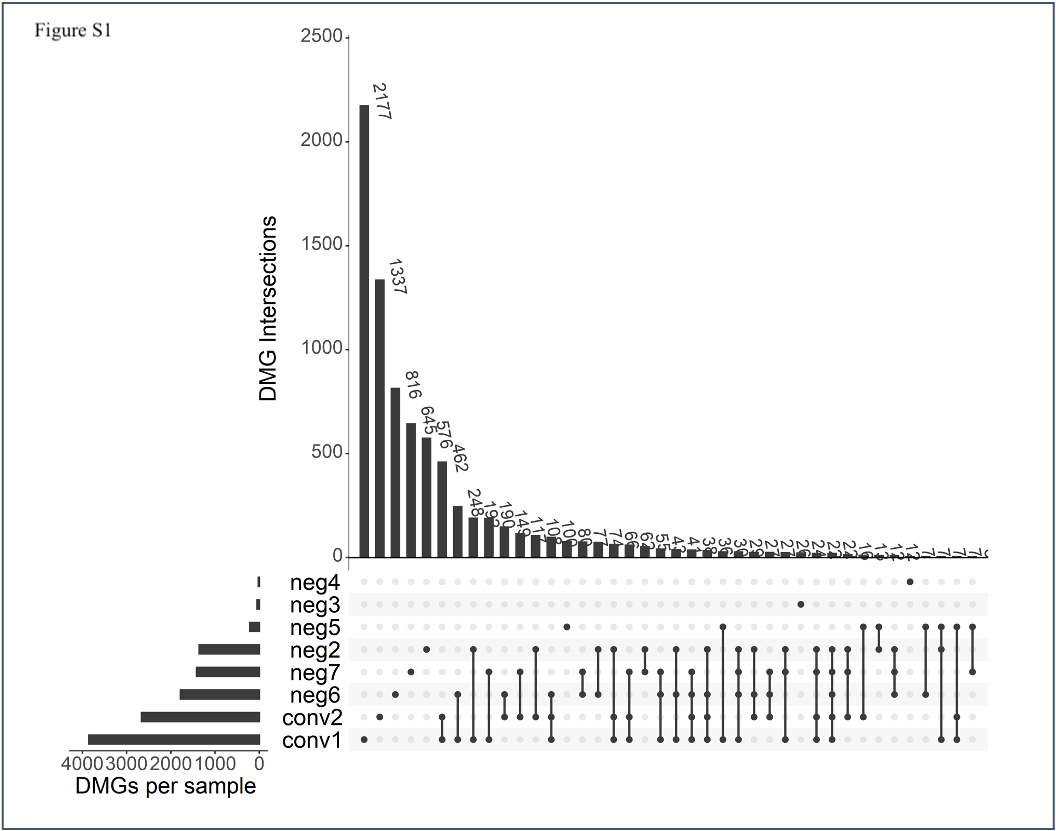
