## Supplementary material for "A Differential DNA Methylome Signature of Pulmonary Immune Cells from Individuals Converting to Latent Tuberculosis Infection": Table S1. IGRA test results.

Table S1. **IGRA test results.** The IGRA results from the study subjects that was IGRA_pos_ at inclusion (0 months) and for the IGRA converters that were IGRA negative at 0 months and IGRA positive at 6 months.

|  | 0 months | | 6 months | |
| --- | --- | --- | --- | --- |
|  | **TB1-Nil** | **TB2-Nil** | **TB1-Nil** | **TB2-Nil** |
| ***Pos 1*** | 0.33 | 0.52 | - | - |
| ***Conv 1*** | 0.03 | 0.06 | 2.43 | 2.28 |
| ***Conv 2*** | -0.01 | 0 | 6.62 | 6.8 |
